# Cardiovascular disease protein biomarkers are associated with kidney function: the Framingham Heart Study

**DOI:** 10.1101/2022.01.04.22268737

**Authors:** Amena Keshawarz, Shih-Jen Hwang, Gha Young Lee, Zhi Yu, Chen Yao, Anna Köttgen, Daniel Levy

## Abstract

**Background:** Biomarkers common to chronic kidney disease (CKD) and cardiovascular disease (CVD) may reflect early impairments underlying both diseases.

**Methods:** We evaluated associations of 71 CVD-related plasma proteins measured in 2,873 Framingham Heart Study (FHS) Offspring cohort participants with cross-sectional continuous eGFR and with longitudinal change in eGFR from baseline to follow-up (ΔeGFR). We also evaluated the associations of the 71 CVD proteins with the following dichotomous secondary outcomes: prevalent CKD stage ≥3 (cross-sectional), new-onset CKD stage ≥3 (longitudinal), and rapid decline in eGFR (longitudinal). Proteins significantly associated with eGFR and ΔeGFR were subsequently validated in 3,951 FHS Third Generation cohort participants and were tested using Mendelian randomization (MR) analysis to infer putatively causal relations between plasma protein biomarkers and kidney function.

**Results:** In cross-sectional analysis, 37 protein biomarkers were significantly associated with eGFR at FDR<0.05 in the FHS Offspring cohort and 20 of these validated in the FHS Third Generation cohort at p<0.05/37. In longitudinal analysis, 27 protein biomarkers were significantly associated with ΔeGFR at FDR<0.05 and 12 of these were validated in the FHS Third Generation cohort at p<0.05/27. Additionally, 35 protein biomarkers were significantly associated with prevalent CKD, five were significantly associated with new-onset CKD, and 17 were significantly associated with rapid decline in eGFR. MR suggested putatively causal relations of melanoma cell adhesion molecule (MCAM; -0.011±0.003 mL/min/1.73m^2^, p=5.11E-5) and epidermal growth factor-containing fibulin-like extracellular matrix protein 1 (EFEMP1; - 0.006±0.002 mL/min/1.73m^2^, p=0.0001) concentration with eGFR.

**Discussion/Conclusions:** Eight protein biomarkers were consistently associated with eGFR in cross-sectional and longitudinal analysis in both cohorts and may capture early kidney impairment; others were implicated in association and causal inference analyses. A subset of CVD protein biomarkers may contribute causally to the pathogenesis of kidney impairment and should be studied as targets for CKD treatment and early prevention.

## Introduction

Chronic kidney disease (CKD) affects approximately 15% of the United States population, including more than 30% of adults over the age of 65 years.^1,2^ Additionally, CKD is among the leading global causes of mortality.^3^ CKD is characterized by kidney damage and impairment of filtration, which can culminate in kidney failure and death.^4^ Kidney function is frequently assessed clinically using estimated glomerular filtration rate (eGFR) based on serum creatinine concentration.^5^ Early stages of CKD are often asymptomatic, and as such, CKD may not be diagnosed until the development of substantial and often irreversible kidney dysfunction. Thus, the identification of biomarkers of subclinical CKD that detect early kidney impairment when treatment is more likely to be beneficial could prove to be important for disease prevention and treatment.

CKD and cardiovascular disease (CVD) share common risk factors including diabetes mellitus (DM) and hypertension,^6^ which are highly prevalent in adults with CKD and are associated with subclinical and clinical CVD risk.^7,8^ More than 50% of CKD cases in the United States are attributable to DM,^9^ and estimates of hypertension prevalence in patients with CKD range between 40% and 60%.^10–12^ Other shared mechanisms contributing to CKD and CVD include inflammation,^13^ activation of the renin-angiotensin-aldosterone system,^13^ oxidative stress,^14,15^ and endothelial dysfunction.^14^ Identifying biomarkers of kidney function may provide insights into shared mechanisms of CKD and CVD.

Given the complex relationship between CKD and CVD, we aimed to identify protein biomarkers of kidney dysfunction and CKD. To this end, we evaluated the associations of 71 CVD-related plasma proteins with CKD traits. These proteins were measured in 7,184 Framingham Heart Study (FHS) participants as a part of the Systems Approach to Biomarker Research in Cardiovascular Disease (SABRe CVD) Initiative.^16^ We tested the cross-sectional and longitudinal associations of the 71 CVD proteins with kidney function traits. We also conducted causal inference analyses using genetic variants associated with these proteins in conjunction with genetic variants from a recently published large GWAS meta-analysis of kidney function traits.^17^

## Materials and Methods

### Discovery sample

The discovery sample for this investigation included participants in the FHS Offspring^18^ cohort who attended the seventh examination cycle (1998-2001; baseline visit). At this visit, serum and plasma samples were collected for measurement of serum creatinine and the 71 CVD-related plasma proteins as part of the SABRe CVD Initiative.^16^ Participants with a baseline eGFR ≥ 60 mL/min/1.73m^2^ who attended the FHS Offspring eighth examination cycle (2005-2008; follow-up visit) where a second serum creatinine measurement was obtained were included in the longitudinal analysis of kidney function.

Participants with a medical record-confirmed diagnosis of heart failure or myocardial infarction at the baseline visit were excluded due to the effect of these diagnoses on biomarker concentrations, yielding a final sample of 2,873 participants for cross-sectional analysis. There were 2,393 participants who attended the follow-up examination and were eligible for inclusion in the longitudinal analyses. All study protocols were approved by the Boston University Medical Center institutional review board, and all study participants provided their informed consent to participate in FHS research investigations. An overview of the study design is presented in **Figure 1**.

**Figure 1.**
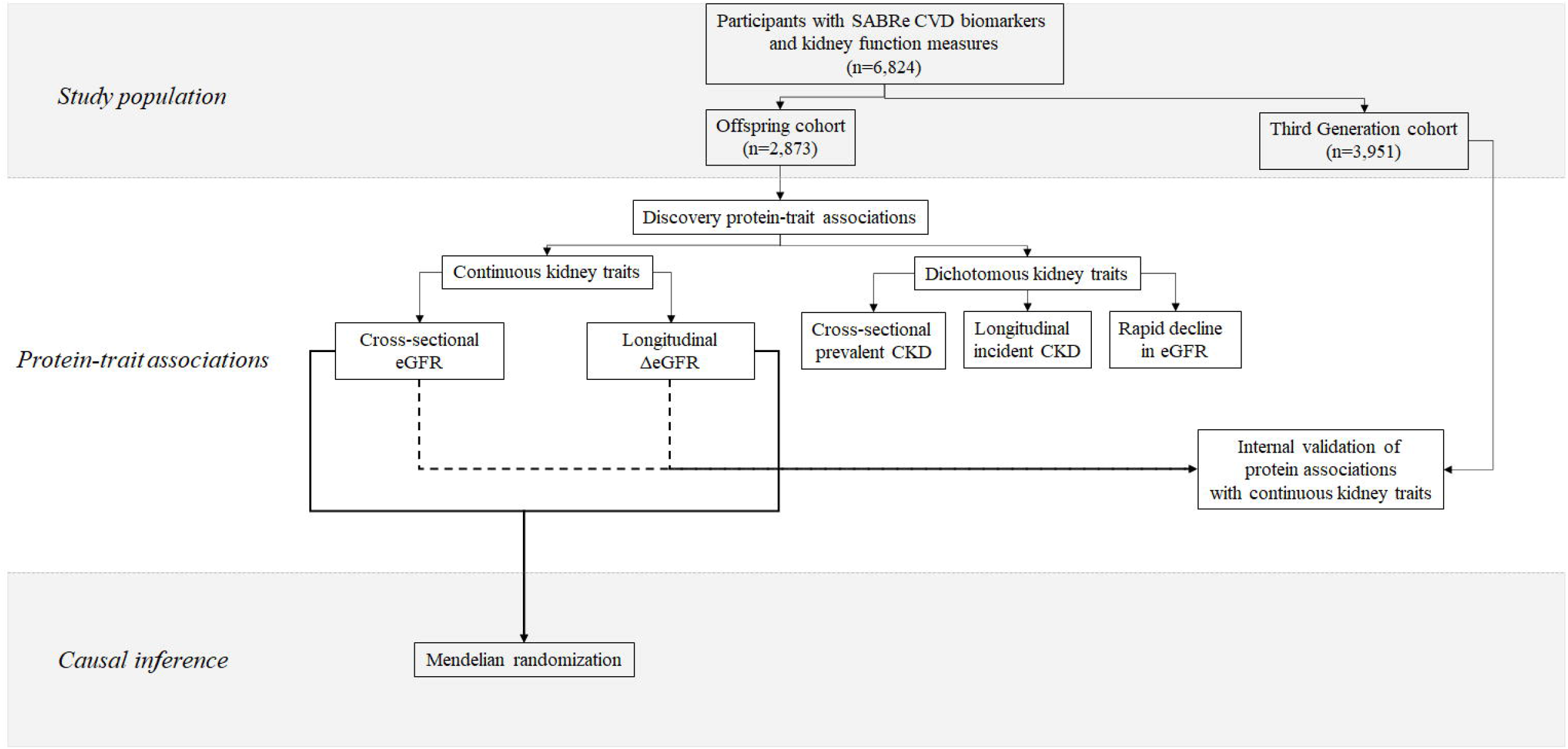
Overview of the study design.

### Clinical examination and definitions

Medical history and fasting blood samples were collected from all study participants at an in-person research center examination.^19^ Body mass index (BMI) was calculated as the ratio of the participant’s weight in kilograms to the square of height in meters. DM status was defined by fasting blood glucose concentration ≥126 mg/dL or use of hypoglycemic medication.^20^ Hypertension was defined by systolic blood pressure ≥ 140 mmHg, diastolic blood pressure ≥ 90 mmHg, or use of antihypertensive medication.^21^ Participants were defined as current smokers if they reported smoking at least one cigarette per day on average during the previous year. Prevalent CVD at the baseline study visit was defined as angina pectoris, coronary insufficiency, cerebrovascular accident, atherothrombotic infarction of brain, transient ischemic attack, cerebral embolism, intracerebral hemorrhage, subarachnoid hemorrhage, or intermittent claudication.

### SABRe CVD biomarkers

Details of measurement of the 71 CVD-related proteins evaluated in this study have been described previously.^16^ Briefly, plasma samples were obtained from all participants at their baseline clinical examinations and stored at -80°C. Samples were assayed and concentrations of 85 biomarkers were quantified using a modified ELISA sandwich approach on a Luminex xMAP platform (Sigma-Aldrich, St. Louis, MO); 71 of these had detectable levels in at least 95% of participant samples and were used in the present analyses. The 71 protein biomarkers used in this study and their molecular weights are presented in **Supplemental Table 1**. Due to the distribution of protein concentration values, inverse-rank normalized protein concentration values were used in all statistical analyses.

### Kidney function traits

Serum creatinine was measured using the modified Jaffe method (Roche Diagnostics, Indianapolis, IN) and calibrated to National Health and Nutrition Examination Survey (NHANES) III creatinine values.^6^ eGFR, an estimate of kidney function, was calculated using the CKD Epidemiology Collaboration (CKD-EPI) creatinine equation.^5^ The annual change in eGFR (ΔeGFR) was calculated for each participant as the follow-up minus baseline eGFR value divided by the number of years between the two visits.

This study primarily focused on the continuous eGFR and ΔeGFR measures of kidney function, with a secondary emphasis on dichotomous traits of CKD and rapid decline in eGFR. Participants were categorized as having CKD stage ≥ 3 at the baseline visit if they had eGFR values <60 mL/min/1.73m^2^. New-onset CKD stage ≥ 3 was defined as a follow-up eGFR <60 mL/min/1.73m^2^ in participants who had eGFR values ≥60 mL/min/1.73m^2^ at baseline and a concomitant ≥30% reduction in eGFR between baseline and follow-up.^22^Rapid decline in eGFR was defined as a decline in eGFR of at least 3 mL/min/1.73m^2^ per year between baseline and follow-up.

### Meta-analyzed GWAS of kidney function traits

Publicly available kidney trait GWAS summary statistics were obtained from the CKDGen Consortium website (https://ckdgen.imbi.uni-freiburg.de/) as reported in a 2019 meta-analysis that identified genetic variants associated with eGFR and CKD (eGFR < 60 mL/min/1.73m^2^) in trans-ethnic and European Ancestry populations.^17^

### Association of protein biomarkers with kidney traits

The primary kidney function outcomes were cross-sectional eGFR at the baseline visit, and longitudinal ΔeGFR (i.e., eGFR at follow-up minus eGFR at baseline). The statistical associations of each protein biomarker with these traits were evaluated using linear mixed models.

The secondary kidney function outcomes evaluated were cross-sectional CKD at the baseline visit, new-onset CKD at follow-up, and longitudinal rapid decline in eGFR. The statistical associations between variation in the protein distribution and these dichotomous kidney function traits were evaluated using logistic regression models.

All multivariable models were adjusted for the following kidney dysfunction risk factors at baseline: age, sex, BMI, systolic blood pressure, total and HDL cholesterol, blood pressure and/or lipid medication use, prevalent CVD, DM, and cigarette smoking status (current smokers vs. non-smokers). Longitudinal analyses (i.e., models evaluating ΔeGFR, new-onset CKD, and rapid decline in eGFR) were additionally adjusted for baseline eGFR, and only those participants with available longitudinal data and who had a baseline eGFR ≥ 60 mL/min/1.73m^2^ were included in the model assessing new-onset CKD (n=2,257). We defined a “clinical model” including all covariates except the protein biomarker of interest (i.e., reduced model), and a “full model” including all covariates in the clinical model and the protein biomarker of interest.

For each protein biomarker, we evaluated the quantitative contribution of the protein biomarker to variation in continuous kidney trait outcomes by comparing the goodness-of-fit R^2^ value of the main and full models. The change in R^2^ (ΔR^2^) illustrates the incremental impact of the addition of the protein biomarker over and above the clinical model. For the dichotomous kidney function traits, we compared receiver operating characteristic (ROC) curves between the clinical and full models using a contrast matrix to quantify the difference in the area under the empirical ROC curve (i.e., the c-statistic). Results of the ROC curve comparison fit a chi-square distribution, which was used to evaluate statistical significance. We calculated the false discovery rate (FDR) for all associations, and FDR values <0.05 were interpreted as statistically significant. Discovery protein-trait statistical analyses were performed in SAS version 9.4 (SAS Institute, Cary, NC).

### Validation of continuous kidney function outcomes

We conducted internal validation of the protein biomarkers significantly associated with eGFR in cross-sectional discovery analysis and with ΔeGFR in longitudinal discovery analyses. Protein biomarkers that were significantly associated with eGFR in the FHS Offspring cohort were subsequently validated in 3,951 participants in the FHS Third Generation cohort^23^ using data from the first (2002-2005) and second (2009-2011) exams of this cohort. Third Generation cohort participants were included in the internal validation sample if they had serum creatinine and the SABRe plasma protein measurements and did not have a medical record-confirmed diagnosis of heart failure or myocardial infarction. Statistical significance of the internal validation analyses was defined as a Bonferroni-corrected p-value of 0.05/total number of proteins included in the validation analysis. Validation analyses were performed in SAS version 9.4 (SAS Institute, Cary, NC).

### Two-sample Mendelian randomization (MR)

We used a two-sample MR approach^24^ to test the hypothesis that protein concentrations associated with eGFR in the discovery and validation protein-trait analyses are causally related to continuous eGFR using data from published kidney trait GWAS.^17^ Prior genome-wide association studies (GWAS) of SABRe proteins in FHS participants identified protein quantitative trait loci (pQTL) variants for 57 of the 71 protein biomarkers.^25^ Of these previously identified variants, we considered only *cis-*pQTL variants for protein biomarkers that were significantly associated with cross-sectional eGFR or longitudinal ΔeGFR in the validation analysis in the Third Generation cohort. *cis-*pQTL variants were defined as those single nucleotide polymorphisms (SNPs) that were located 1 megabase upstream or downstream of the protein-coding gene’s transcription start site. Furthermore, *cis*-pQTL variants were pruned at linkage disequilibrium r^2^ < 0.01, and only those that overlapped with SNPs from the kidney function GWAS were used as instrumental variables for the protein biomarkers.

For protein biomarkers with only one pQTL variant as an instrumental variant in MR analysis after pruning, the causal effect was calculated using the Wald test. For protein biomarkers with more than one pQTL variant after pruning, the causal effect was calculated using inverse-variance weighted meta-analyzed estimates. MR results were interpreted as statistically significant after applying a Bonferroni correction for the total number of unique protein biomarkers tested across all outcomes (i.e., p<0.05/n, where n is the number of protein-trait associations tested).

In the case of multiple *cis*-pQTL variants contributing to a significant weighted causal estimate, we conducted sensitivity analyses for heterogeneity. When more than two pQTL variants or SNPs contributed to a significant weighted causal estimate, we additionally tested for horizontal pleiotropy and evaluated the change in the effect estimate after excluding one variant at a time from the calculation (i.e., leave-one-out analysis).

All MR analyses were conducted in R version 4.0.2 using the *TwoSampleMR* package.^24,26^

## Results

### Participant characteristics

Baseline characteristics of participants in the FHS Offspring and Third Generation cross-sectional cohorts are presented in **Table 1**. The mean age of participants in the Offspring cohort was 60 ± 9 years and mean eGFR was 84 ± 16 mL/min/1.73m^2^. Seven percent of the Offspring cohort had CKD at baseline, while the prevalence of DM and hypertension were 10% and 43%, respectively. The mean age of participants in the Third Generation cohort was 40 ± 9 years and mean eGFR was 102 ± 14 mL/min/1.73m^2^. Less than one percent of the Third Generation cohort sample had CKD, while the prevalence of DM and hypertension were 3% and 18%, respectively.

**Table 1.** Baseline demographic and clinical participant characteristics.

| Characteristic | Offspring cohort<br>(n=2,873) | Third Generation cohort<br>(n=3,951) |
| --- | --- | --- |
| Age, years | 60 ± 9 | 40 ± 9 |
| Women, n (%) | 1,573 (55%) | 2,109 (53%) |
| eGFR, mL/min/1.73m<sup>2</sup> | 84 ± 16 | 102 ± 14 |
| CKD, n (%) | 210 (7%) | 10 (0.3%) |
| BMI, kg/m<sup>2</sup> | 27.95 ± 5.22 | 26.87 ± 5.55 |
| Fasting glucose, mg/dL | 102.99 ± 24.59 | 95.02 ± 17.9 |
| Systolic blood pressure, mmHg | 126.18 ± 18.54 | 116.69 ± 14.02 |
| Diastolic blood pressure, mmHg | 74.15 ± 9.7 | 75.26 ± 9.63 |
| Total cholesterol, mg/dL | 200.64 ± 36.42 | 188.81 ± 35.46 |
| HDL cholesterol, mg/dL | 54.41 ± 17.2 | 54.41 ± 16.16 |
| DM, n (%) | 280 (10%) | 114 (3%) |
| Hypertension, n (%) | 1242 (43%) | 726 (18%) |
| Current cigarette smoker, n (%) | 362 (13%) | 605 (15%) |
| Cholesterol-lowering medication use, n (%) | 564 (20%) | 341 (9%) |
| Prevalent cardiovascular disease, n (%) | 308 (11%) | 38 (1%) |

Abbreviations: eGFR, estimated glomerular filtration rate; CKD, chronic kidney disease; BMI, body mass index;
HDL, high density lipoprotein; DM, diabetes mellitus
All continuous characteristics are presented as mean ± standard deviation. All categorical variables are presented as total n (column %) in that category.

**Table 2.**
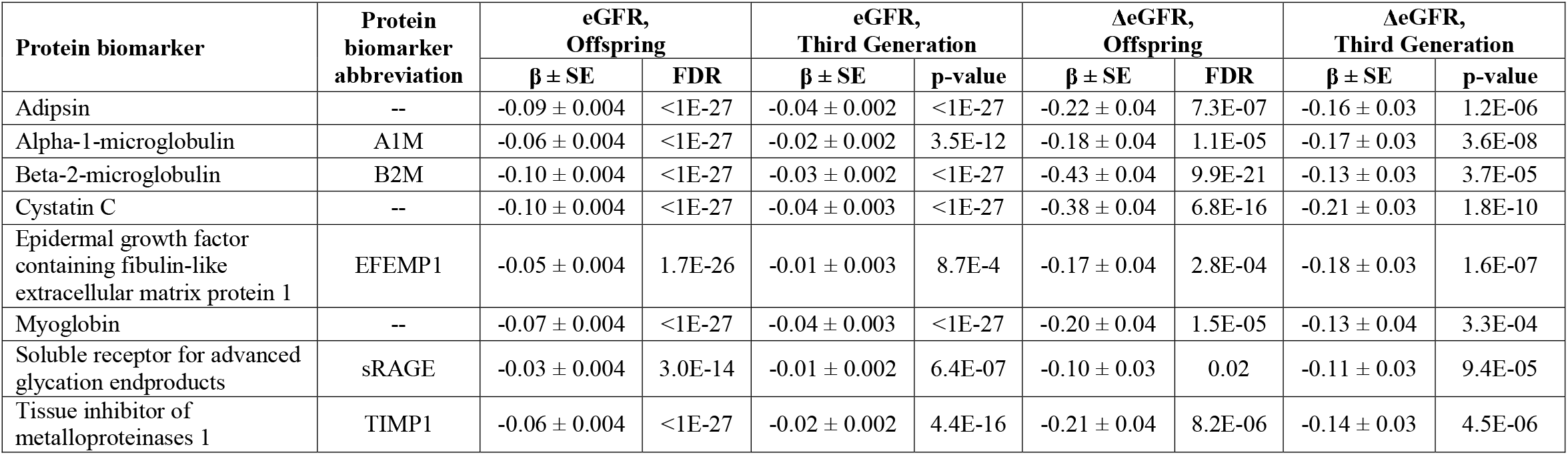
Biomarkers associated with eGFR and ΔeGFR in the FHS Offspring and Third Generation cohorts.

Follow-up characteristics of FHS Offspring (discovery) cohort participants included in longitudinal analysis of ΔeGFR, new-onset CKD, and rapid decline in eGFR are presented in **Supplemental Table 2**. The mean age of Offspring cohort participants at follow-up was 66 ± 9 years and mean annual ΔeGFR between baseline and follow-up was -0.97 ± 1.73 mL/min/1.73m^2^. Among participants who did not have CKD at baseline, 3% experienced new-onset CKD during follow-up while 91% maintained their eGFR at over 60 mL/min/1.73m^2^. The remaining participants had a follow-up eGFR < 60 mL/min/1.73m^2^, but had a reduction in eGFR < 30%. Eleven percent of the Offspring cohort sample included in longitudinal analysis experienced rapid decline in eGFR between baseline and follow-up.

### Discovery and validation of cross-sectional and longitudinal protein-eGFR associations

Cross-sectional associations of each of the 71 protein biomarkers with eGFR and the incremental increase in model R^2^ associated with each protein biomarker are presented in **Supplemental Table 3**. Thirty-seven protein biomarkers were significantly associated with eGFR (at FDR<0.05) in cross-sectional analysis, and of these, all but one (butyrylcholine esterase) had an inverse association with eGFR. Six protein biomarkers were significantly associated with a change in R^2^ ≥ 5%, including alpha-1-microglobulin (A1M), adipsin, beta-2-microglobulin (B2M), cystatin C, myoglobin, and resistin. Of the 37 protein biomarkers that were significantly associated with eGFR in the Offspring cohort in cross-sectional analysis, 20 were also significantly associated with eGFR in the Third Generation cohort at p=0.05/37=0.00135 **(Supplemental Table 4)**.

Longitudinal associations of each of the 71 protein biomarkers with ΔeGFR and the incremental increase in model R^2^ associated with each protein biomarker are presented in **Supplemental Table 5**. Twenty-seven protein biomarkers were significantly associated with ΔeGFR at FDR<0.05. All significant protein biomarkers except two (plasminogen activator inhibitor 1 [PAI1] and dipeptidyl dipeptidase [DPP4]) were inversely associated with ΔeGFR. B2M and cystatin C were associated with the largest increases in R^2^ (ΔR^2^ = 3.3% and 2.5%, respectively). Of the 27 proteins that were significantly associated with ΔeGFR in the Offspring cohort in longitudinal analysis, 12 were also significantly associated with ΔeGFR in the Third Generation cohort at p=0.05/27=0.00185 (**Supplemental Table 6)**.

### Discovery protein-trait associations: Cross-sectional associations with secondary outcomes in the Offspring cohort

Cross-sectional associations of each of the 71 protein biomarkers with prevalent CKD and the associated change in the C-statistic are presented in **Supplemental Table 7**. Thirty-five protein biomarkers were significantly associated with prevalent CKD at FDR<0.05, and of these, all but one (angiopoietin-like 3) were associated with greater log-odds of CKD. Ten protein biomarkers were associated with a statistically significant change in the C-statistic.

### Discovery protein-trait associations: Longitudinal associations with secondary outcomes in the Offspring cohort

Five protein biomarkers were significantly associated with higher log-odds of new-onset CKD at FDR<0.05 (**Supplemental Table 8)**, while 17 protein biomarkers were significantly associated with higher log-odds of rapid decline in eGFR **(Supplemental Table 9)**. No biomarkers were associated with a significant increase in the model C-statistic for either rapid decline in eGFR or new-onset CKD.

### Causal inference

Twenty-four unique proteins were significantly associated with eGFR in both the Offspring and Third Generation in cross-sectional and/or longitudinal analyses; therefore, statistical significance for all MR analyses was set at a Bonferroni-corrected p-value of 0.05/24 = 0.00208. The 24 protein biomarkers associated with eGFR had a total of 25 *cis-*pQTL variants that overlapped with SNPs in the eGFR GWAS^17^ and were included in MR analyses. Significant results of MR analysis (protein → kidney trait) for proteins that were significantly associated with eGFR or with ΔeGFR are presented in **Table 3**; full MR results for all protein biomarkers with *cis*-pQTL variants are presented in **Supplemental Table 10**. Higher concentrations of epidermal growth factor-containing fibulin-like extracellular matrix protein 1 (EFEMP1) and melanoma cell adhesion molecule (MCAM) were significantly associated with lower eGFR (β ± SE -0.0064 ± 0.0016 and -0.0111 ± 0.0027, respectively), consistent with a potentially causal relationship. Sensitivity analysis of the two EFEMP1 pQTL variants indicated no heterogeneity (p=0.69, **Supplemental Table 9**).

**Table 3.** Causal inference results: MR results for biomarkers significantly associated with eGFR in cross-sectional analyses.

| Protein biomarker exposure | Protein biomarker abbreviation | Kidney trait outcome | n<sub>snp</sub> | $\beta \pm SE$ | p-value |
| --- | --- | --- | --- | --- | --- |
| Melanoma cell adhesion molecule | MCAM | eGFR (continuous) | 1 | -0.0111 ± 0.0027 | 5.11E-5 |
| Epidermal growth factor containing fibulin-like extracellular matrix protein 1 | EFEMP1 | eGFR (continuous) | 2 | -0.0064 ± 0.0016 | 1.04E-4 |

## Discussion/Conclusion

We identified a total of 43 CVD-related protein biomarkers associated with eGFR, a continuous measure of kidney function, in cross-sectional and/or longitudinal discovery analysis. Among these, eight proteins were significantly associated with both eGFR and ΔeGFR in the FHS Offspring discovery sample and the FHS Third Generation validation sample. Of the 37 proteins associated with eGFR and 27 proteins associated with ΔeGFR in the FHS Offspring cohort (i.e. discovery), 20 and 12 were associated with the corresponding trait in the FHS Third Generation cohort (i.e. validation). This group of protein biomarkers included EFEMP1 and MCAM, which were putatively causally related to eGFR in MR analysis. To our knowledge, causal relations of protein biomarkers to CKD based on an integrative genomic approach have not been reported previously.

CKD and CVD share several risk factors; identifying early mechanisms linking the two disease processes may provide insights into targeted interventions to prevent or delay the onset of clinically overt disease. The most notable shared risk factors are DM and hypertension. DM is associated with systemic inflammation and oxidative stress,^27^ which in turn are associated with microvascular and macrovascular complications and kidney damage.^28,29^ Hypertension results in prolonged exposure of the heart, kidney, and vasculature to elevated hemodynamic load that can contribute to left ventricular hypertrophy, vascular stiffening, heart failure, and CKD.^30^ Over-activation of the renin-angiotensin-aldosterone system, which increases blood pressure through multiple mechanisms, can damage glomerular cells.^13^ Treatment of hypertension can prevent CKD progression,^31^ while conversely, kidney disease can lead to resistant hypertension.^32^ Furthermore, inflammation also contributes to development of both CKD and CVD. In CKD, the concentration of inflammatory proteins is inversely related to eGFR, and the cytokine interleukin (IL)-6 is a reliable predictor of adverse clinical outcomes in individuals with CKD.^14,33,34^ Chronic inflammation also may cause oxidative stress and endothelial dysfunction, which contribute to microvascular damage, new-onset or worsening CKD, and clinical CVD.^15^

The eGFR values observed in the FHS Offspring participants included in the discovery stage of this study were mostly in the normal range and few individuals experience a rapid decline in eGFR (n=265; 11%) or progressed to new-onset CKD (n=72; 3%) during follow-up. Under normal conditions, the glomerulus is able to freely filter proteins with molecular weight (MW) < 70 kilodaltons (kDa), and as such, many of the proteins identified in the cross-sectional and longitudinal protein-trait analyses may reflect early impaired kidney clearance of proteins with low molecular weights with a resultant increase in concentrations of circulating proteins.^35^ Indeed, the vast majority of protein biomarkers showed negative associations with eGFR (i.e., lower eGFR was associated with higher protein levels), thus many of the protein biomarkers identified in this study are likely downstream biomarkers of reduced kidney function. Accordingly, the protein biomarkers with consistent and highly significant associations are well-established markers of kidney function, and our validation in the Third Generation cohort of many of the discovery associations observed in the Offspring cohort emphasize associations of these proteins with kidney function in a younger, healthier population with little prevalent CKD. Eight protein biomarkers were associated with both eGFR and ΔeGFR in both the Offspring and Third Generation cohorts, including adipsin, adrenomedullin (ADM), A1M, B2M, collagen type XVIII alpha 1 (COL18A1), cystatin C, EFEMP1, and fibroblast growth factor 23 (FGF23). Of these, cystatin C (MW 15.8 kDa) and B2M (MW 13.7 kDa) were associated with all kidney traits and are used in alternative eGFR calculations due to their low MW and their role as early indicators of kidney impairment, and they are additionally associated with CVD outcomes.^16,36–39^ Other protein biomarkers significantly associated with eGFR and ΔeGFR are also associated with inflammation and kidney disease and may represent non-traditional markers of kidney function due to parallel mechanisms relevant also to CVD. A1M (MW 29.8 kDa), while having a slightly higher MW than either cystatin C or B2M, has also been proposed as a biomarker of kidney disease progression.^40–42^ Similarly, adipsin (MW 27.0 kDa), ADM (MW 20.4 kDa), FGF23 (MW 28.0 kDa), and endostatin, a 20 kDa fragment of the 178.2 kDa COL18A1, were all reported to be elevated in the setting of kidney disease in prior studies. ^43–47^ These protein biomarkers showed a robust association with kidney traits and it is possible that they reflect CKD risk early in the subclinical disease process.

We hypothesized *a priori* that a subset of proteins associated with kidney function may be causal contributors to renal impairment. A recent MR analysis suggested a putatively causal role of melanoma-derived growth regulatory protein (MIA), cystatin M, and carbonic anhydrase III in kidney disease.^55^ While these proteins were not included in our assay, our MR analysis evaluated 24 new protein biomarkers and revealed putatively causal inverse relations of EFEMP1 and MCAM with eGFR. EFEMP1 is an extracellular matrix protein involved in both cellular structure and signaling, and it was associated with worse eGFR cross-sectionally and with ΔeGFR longitudinally, with prevalent CKD, and with rapid decline in eGFR. A prior study reported elevated EFEMP1 concentration in human kidney tissue with immunoglobulin A (IgA) nephropathy.^56^ The causal mechanism is still unclear, but animal models suggest that EFEMP1 may be associated with vascular remodeling in hypertension.^57^ MCAM, also known as MUC18 or CD146, is expressed in endothelial cells and is significantly elevated in individuals with diabetic nephropathy^58,59^ and CKD.^60,61^ Results of our protein-trait analysis similarly showed an association between MCAM with both eGFR and CKD in cross-sectional analyses.

This study has several limitations that must be noted. The primary limitation is that a custom assay used to measure the protein biomarkers in this study, which limited external replication of our results. To address this limitation, we performed validation of the discovery results from the FHS Offspring cohort in participants from the younger and healthier Third Generation cohort in whom CKD was rare (0.3% prevalent CKD at baseline). Independent external replication of the protein biomarkers identified in this study is needed.

In addition, we estimated kidney function based on a single creatinine measurement at both baseline and follow-up, rather than using multiple eGFR values at each time point. As such, our definition of CKD stage ≥ 3 does not mirror the recommended clinical diagnosis of CKD, which requires the presence of kidney structural or functional abnormalities for at least three months.^62^ Furthermore, the younger and healthier cohort used for validation analyses limited our ability to identify protein biomarkers of changing kidney function and CKD in longitudinal analysis. Albuminuria and blood urea nitrogen may be clinically useful in the evaluation of patients with impaired kidney function, but these data were not available for analysis in this study. Our study population was limited to adults of European ancestry, which may limit generalizability to other ancestry groups. Functional studies of the proteins reported to have putatively causal relations to kidney function in this investigation may be warranted.

The primary strengths of this study are the large sample size and the integrative genomic approach we employed. We reported cross-sectional and longitudinal protein-trait association results to identify numerous proteins associated with kidney function and its change over time. By integrating pQTL data for the proteins we measured with genetic variants from large GWAS of kidney disease traits we identified putatively causal proteins for kidney function that represent promising candidates for further studies with the ultimate goal to better treat or prevent CKD.

The results of the comprehensive cross-sectional and longitudinal analyses in this study validate proteins that may detect of impaired kidney function early in the disease process when treatment is most likely to be beneficial. We identified robust, significant associations between 20 protein biomarkers with eGFR in cross-sectional discovery and validation analyses and 12 proteins with ΔeGFR in longitudinal analyses both in discovery and validation. Additionally, two proteins were found to be putatively causal for reduced kidney function in causal inference testing. Further studies are necessary to determine if any of the proteins identified by MR can serve as useful biomarkers for CKD either individually or in combination with known and validated biomarkers.

## Data Availability

All data produced in the present study are either publicly available or available upon request through the Database of Genotypes and Phenotypes.

## Statements

### Statement of ethics

The study protocol was reviewed and approved by the Boston University Medical Center Institutional Review Board. All study participants provided their written informed consent for their data and biological samples to be used.

### Disclaimer

The views expressed in this manuscript are those of the authors and do not necessarily represent the views of the National Heart, Lung, and Blood Institute; the National Institutes of Health; or the U.S. Department of Health and Human Services.

### Author contributions

A Keshawarz, GY Lee, S Hwang, Z Yu, and C Yao contributed to data analysis. All authors contributed to data interpretation. GY Lee, S Hwang, A Köttgen, and D Levy conceived the research study. A Keshawarz drafted the manuscript. GY Lee, S Hwang, C Yao, A Köttgen, and D Levy provided critical revisions. All authors gave final approval for publication and are the guarantors of this work.

### Data availability statement

The data presented in these analyses can be accessed through the National Center for Biotechnology Information Database of Genotypes and Phenotypes (accession number, phs00007.v29.p10).

## References

1. Saran R, Robinson B, Abbott KC, et al. US Renal Data System 2017 Annual Data Report: Epidemiology of Kidney Disease in the United States. Am J kidney Dis Off J Natl Kidney Found. 2018;71(3 Suppl 1):A7. doi:10.1053/j.ajkd.2018.01.002

2. Joubert BR, Felix JF, Yousefi P, et al. DNA Methylation in Newborns and Maternal Smoking in Pregnancy: Genome-wide Consortium Meta-analysis. Am J Hum Genet. 2016;98(4):680–696. doi:https://doi.org/10.1016/j.ajhg.2016.02.019

3. Bikbov B, Purcell CA, Levey AS, et al. Global, regional, and national burden of chronic kidney disease, 1990–2017: a systematic analysis for the Global Burden of Disease Study 2017. Lancet. 2020;395(10225):709–733. doi:10.1016/S0140-6736(20)30045-3

4. Gansevoort RT, Matsushita K, van der Velde M, et al. Lower estimated GFR and higher albuminuria are associated with adverse kidney outcomes. A collaborative meta-analysis of general and high-risk population cohorts. Kidney Int. 2011;80(1):93–104. doi:10.1038/ki.2010.531

5. Levey AS, Stevens LA, Schmid CH, et al. A new equation to estimate glomerular filtration rate. Ann Intern Med. 2009;150(9):604–612. doi:10.7326/0003-4819-150-9-200905050-00006

6. Fox CS, Larson MG, Leip EP, Culleton B, Wilson PWF, Levy D. Predictors of New-Onset Kidney Disease in a Community-Based Population. JAMA. 2004;291(7):844–850. doi:10.1001/jama.291.7.844

7. Flint AC, Conell C, Ren X, et al. Effect of Systolic and Diastolic Blood Pressure on Cardiovascular Outcomes. N Engl J Med. 2019;381(3):243–251. doi:10.1056/NEJMoa1803180

8. Dokken BB. The Pathophysiology of Cardiovascular Disease and Diabetes: Beyond Blood Pressure and Lipids. Diabetes Spectr. 2008;21(3):160LP–165. doi:10.2337/diaspect.21.3.160

9. Zelnick LR, Weiss NS, Kestenbaum BR, et al. Diabetes and CKD in the United States Population, 2009–2014. Clin J Am Soc Nephrol. 2017;12(12):1984LP–1990. doi:10.2215/CJN.03700417

10. United States Renal Data System. 2020 USRDS Annual Data Report: Epidemiology of Kidney Disease in the United States.; 2020.

11. Kibria GM Al, Crispen R. Prevalence and trends of chronic kidney disease and its risk factors among US adults: An analysis of NHANES 2003-18. Prev Med reports. 2020;20:101193. doi:10.1016/j.pmedr.2020.101193

12. Centers for Disease Control and Prevention. Chronic Kidney Disease Surveillance System—United States. Published 2020. http://www.cdc.gov/ckd

13. Gajjala PR, Sanati M, Jankowski J. Cellular and Molecular Mechanisms of Chronic Kidney Disease with Diabetes Mellitus and Cardiovascular Diseases as Its Comorbidities. Front Immunol. 2015;6:340. doi:10.3389/fimmu.2015.00340

14. Cachofeiro V, Goicochea M, de Vinuesa SG, Oubiña P, Lahera V, Luño J. Oxidative stress and inflammation, a link between chronic kidney disease and cardiovascular disease: New strategies to prevent cardiovascular risk in chronic kidney disease. Kidney Int. 2008;74:S4–S9. doi:10.1038/ki.2008.516

15. Podkowińska A, Formanowicz D. Chronic Kidney Disease as Oxidative Stress-and Inflammatory-Mediated Cardiovascular Disease. Antioxidants (Basel, Switzerland). 2020;9(8):752. doi:10.3390/antiox9080752

16. Ho JE, Lyass A, Courchesne P, et al. Protein Biomarkers of Cardiovascular Disease and Mortality in the Community. J Am Heart Assoc. 2018;7(14):e008108. doi:10.1161/JAHA.117.008108

17. Wuttke M, Li Y, Li M, et al. A catalog of genetic loci associated with kidney function from analyses of a million individuals. Nat Genet. 2019;51(6):957–972. doi:10.1038/s41588-019-0407-x

18. Kannel WB, Feinleib M, McNamara PM, Garrison RJ, Castelli WP. An investigation of coronary heart disease in families: the Framinham Offspring Study. Am J Epidemiol. 1979;110(3):281–290. doi:10.1093/oxfordjournals.aje.a112813

19. Tsao CW, Vasan RS. Cohort Profile: The Framingham Heart Study (FHS): overview of milestones in cardiovascular epidemiology. Int J Epidemiol. 2015;44(6):1800–1813. doi:10.1093/ije/dyv337

20. Association AD. 2. Classification and Diagnosis of Diabetes: <Standards of Medical Care in Diabetes—2020>; Diabetes Care. 2020;43(Supplement 1):S14 LP–S31. doi:10.2337/dc20-S002

21. Chobanian A V, Bakris GL, Black HR, et al. Seventh Report of the Joint National Committee on Prevention, Detection, Evaluation, and Treatment of High Blood Pressure. Hypertension. 2003;42(6):1206–1252. doi:10.1161/01.HYP.0000107251.49515.c2

22. Levey AS, Inker LA, Matsushita K, et al. GFR decline as an end point for clinical trials in CKD: a scientific workshop sponsored by the National Kidney Foundation and the US Food and Drug Administration. Am J kidney Dis Off J Natl Kidney Found. 2014;64(6):821–835. doi:10.1053/j.ajkd.2014.07.030

23. Splansky GL, Corey D, Yang Q, et al. The Third Generation Cohort of the National Heart, Lung, and Blood Institute’s Framingham Heart Study: Design, Recruitment, and Initial Examination. Am J Epidemiol. 2007;165(11):1328–1335. doi:10.1093/aje/kwm021

24. Hemani G, Zheng J, Elsworth B, et al. The MR-Base platform supports systematic causal inference across the human phenome. Elife. 2018;7. doi:10.7554/eLife.34408

25. Yao C, Chen G, Song C, et al. GenomeLwide mapping of plasma protein QTLs identifies putatively causal genes and pathways for cardiovascular disease. Nat Commun. 2018;9(1):3268. doi:10.1038/s41467-018-05512-x

26. Hemani G, Tilling K, Davey Smith G. Orienting the causal relationship between imprecisely measured traits using GWAS summary data. PLoS Genet. 2017;13(11):e1007081. doi:10.1371/journal.pgen.1007081

27. Odegaard AO, Jacobs DRJ, Sanchez OA, Goff DCJ, Reiner AP, Gross MD. Oxidative stress, inflammation, endothelial dysfunction and incidence of type 2 diabetes. Cardiovasc Diabetol. 2016;15:51. doi:10.1186/s12933-016-0369-6

28. Lv W, Booz GW, Wang Y, Fan F, Roman RJ. Inflammation and renal fibrosis: Recent developments on key signaling molecules as potential therapeutic targets. Eur J Pharmacol. 2018;820:65–76. doi:10.1016/j.ejphar.2017.12.016

29. Jha JC, Ho F, Dan C, Jandeleit-Dahm K. A causal link between oxidative stress and inflammation in cardiovascular and renal complications of diabetes. Clin Sci (Lond). 2018;132(16):1811–1836. doi:10.1042/CS20171459

30. Nwabuo CC, Appiah D, Moreira HT, et al. Long-term cumulative blood pressure in young adults and incident heart failure, coronary heart disease, stroke, and cardiovascular disease: The CARDIA study. Eur J Prev Cardiol. Published online April 2020:2047487320915342. doi:10.1177/2047487320915342

31. Ptinopoulou AG, Pikilidou MI, Lasaridis AN. The effect of antihypertensive drugs on chronic kidney disease: a comprehensive review. Hypertens Res. 2013;36(2):91–101. doi:10.1038/hr.2012.157

32. Carey RM, Calhoun DA, Bakris GL, et al. Resistant Hypertension: Detection, Evaluation, and Management: A Scientific Statement From the American Heart Association. Hypertens (Dallas, Tex 1979). 2018;72(5):e53–e90. doi:10.1161/HYP.0000000000000084

33. Sun J, Axelsson J, Machowska A, et al. Biomarkers of Cardiovascular Disease and Mortality Risk in Patients with Advanced CKD. Clin J Am Soc Nephrol. 2016;11(7):1163–1172. doi:10.2215/CJN.10441015

34. Gupta J, Mitra N, Kanetsky PA, et al. Association between albuminuria, kidney function, and inflammatory biomarker profile in CKD in CRIC. Clin J Am Soc Nephrol. 2012;7(12):1938–1946. doi:10.2215/CJN.03500412

35. Strober W, Waldmann TA. The role of the kidney in the metabolism of plasma proteins. Nephron. 1974;13(1):35–66. doi:10.1159/000180368

36. Argyropoulos CP, Chen SS, Ng Y-H, et al. Rediscovering Beta-2 Microglobulin As a Biomarker across the Spectrum of Kidney Diseases. Front Med. 2017;4:73. doi:10.3389/fmed.2017.00073

37. Joseph Y, N. Be, C. Ma, et al. Impact of Kidney Function on the Blood Proteome and on Protein Cardiovascular Risk Biomarkers in Patients With Stable Coronary Heart Disease. J Am Heart Assoc. 2020;9(15):e016463. doi:10.1161/JAHA.120.016463

38. Liu X, Foster MC, Tighiouart H, et al. Non-GFR Determinants of Low-Molecular-Weight Serum Protein Filtration Markers in CKD. Am J kidney Dis Off J Natl Kidney Found. 2016;68(6):892–900. doi:10.1053/j.ajkd.2016.07.021

39. Christensson A, Ash JA, DeLisle RK, et al. The Impact of the Glomerular Filtration Rate on the Human Plasma Proteome. PROTEOMICS – Clin Appl. 2018;12(3):1700067. doi:https://doi.org/10.1002/prca.201700067

40. Luczak M, Formanowicz D, Pawliczak E, Wanic-Kossowska M, Wykretowicz A, Figlerowicz M. Chronic kidney disease-related atherosclerosis - proteomic studies of blood plasma. Proteome Sci. 2011;9:25. doi:10.1186/1477-5956-9-25

41. Jotwani V, Katz R, Ix JH, et al. Urinary Biomarkers of Kidney Tubular Damage and Risk of Cardiovascular Disease and Mortality in Elders. Am J Kidney Dis. 2018;72(2):205–213. doi:10.1053/j.ajkd.2017.12.013

42. Vyssoulis GP, Tousoulis D, Antoniades C, Dimitrakopoulos S, Zervoudaki A, Stefanadis C. α-1 Microglobulin as a New Inflammatory Marker in Newly Diagnosed Hypertensive Patients:. Am J Hypertens. 2007;20(9):1016–1021. doi:10.1016/j.amjhyper.2007.01.010

43. Volanakis JE, Barnum SR, Giddens M, Galla JH. Renal Filtration and Catabolism of Complement Protein D. N Engl J Med. 1985;312(7):395–399. doi:10.1056/NEJM198502143120702

44. Velho G, Ragot S, Mohammedi K, et al. Plasma Adrenomedullin and Allelic Variation in the ADM Gene and Kidney Disease in People With Type 2 Diabetes. Diabetes. 2015;64(9):3262–3272. doi:10.2337/db14-1852

45. Fliser D, Kollerits B, Neyer U, et al. Fibroblast Growth Factor 23 (FGF23) Predicts Progression of Chronic Kidney Disease: The Mild to Moderate Kidney Disease (MMKD) Study. J Am Soc Nephrol. 2007;18(9):2600LP–2608. doi:10.1681/ASN.2006080936

46. Dieplinger B, Mueller T, Kollerits B, et al. Pro-A-type natriuretic peptide and pro-adrenomedullin predict progression of chronic kidney disease: the MMKD Study. Kidney Int. 2009;75(4):408–414. doi:10.1038/ki.2008.560

47. Pena MJ, Heinzel A, Heinze G, et al. A panel of novel biomarkers representing different disease pathways improves prediction of renal function decline in type 2 diabetes. PLoS One. 2015;10(5):e0120995–e0120995. doi:10.1371/journal.pone.0120995

48. Lau ES, Paniagua SM, Zarbafian S, et al. Cardiovascular Biomarkers of Obesity and Overlap With Cardiometabolic Dysfunction. J Am Heart Assoc. 2021;10(14):e020215–e020215. doi:10.1161/JAHA.120.020215

49. Lampón N, Hermida-Cadahia EF, Riveiro A, Tutor JC. Association between butyrylcholinesterase activity and low-grade systemic inflammation. Ann Hepatol. 2012;11(3):356–363.

50. Małgorzewicz S, Skrzypczak-Jankun E, Jankun J. Plasminogen activator inhibitor-1 in kidney pathology (Review). Int J Mol Med. 2013;31(3):503–510. doi:10.3892/ijmm.2013.1234

51. Eddy AA, Fogo AB. Plasminogen Activator Inhibitor-1 in Chronic Kidney Disease: Evidence and Mechanisms of Action. J Am Soc Nephrol. 2006;17(11):2999LP–3012. doi:10.1681/ASN.2006050503

52. Kanasaki K. The role of renal dipeptidyl peptidase-4 in kidney disease: renal effects of dipeptidyl peptidase-4 inhibitors with a focus on linagliptin. Clin Sci (Lond). 2018;132(4):489–507. doi:10.1042/CS20180031

53. Ma L-J, Fogo AB. PAI-1 and kidney fibrosis. Front Biosci (Landmark Ed. 2009;14:2028–2041. doi:10.2741/3361

54. Shi S, Koya D, Kanasaki K. Dipeptidyl peptidase-4 and kidney fibrosis in diabetes. Fibrogenesis Tissue Repair. 2016;9(1):1. doi:10.1186/s13069-016-0038-0

55. Matías-García PR, Wilson R, Guo Q, et al. Plasma Proteomics of Renal Function: A Transethnic Meta-Analysis and Mendelian Randomization Study. J Am Soc Nephrol. 2021;32(7):1747LP–1763. doi:10.1681/ASN.2020071070

56. Paunas FTI, Finne K, Leh S, et al. Characterization of glomerular extracellular matrix in IgA nephropathy by proteomic analysis of laser-captured microdissected glomeruli. BMC Nephrol. 2019;20(1):410. doi:10.1186/s12882-019-1598-1

57. Lin Z, Wang Z, Li G, Li B, Xie W, Xiang D. Fibulin-3 may improve vascular health through inhibition of MMP-2/9 and oxidative stress in spontaneously hypertensive rats. Mol Med Rep. 2016;13(5):3805–3812. doi:10.3892/mmr.2016.5036

58. Fan Y, Fei Y, Zheng L, et al. Expression of Endothelial Cell Injury Marker Cd146 Correlates with Disease Severity and Predicts the Renal Outcomes in Patients with Diabetic Nephropathy. Cell Physiol Biochem. 2018;48(1):63–74. doi:10.1159/000491663

59. Saito T, Saito O, Kawano T, et al. Elevation of serum adiponectin and CD146 levels in diabetic nephropathy. Diabetes Res Clin Pract. 2007;78(1):85–92. doi:10.1016/j.diabres.2007.02.014

60. Bardin N, Moal V, Anfosso F, et al. Soluble CD146, a novel endothelial marker, is increased in physiopathological settings linked to endothelial junctional alteration. Thromb Haemost. 2003;90(5):915–920. doi:10.1160/TH02-11-0285

61. Wang N, Fan Y, Ni P, et al. High glucose effect on the role of CD146 in human proximal tubular epithelial cells in vitro. J Nephrol. 2008;21(6):931–940.

62. Levin A, Stevens P, Bilous RW, et al. Kidney disease: Improving global outcomes (KDIGO) CKD work group. KDIGO 2012 clinical practice guideline for the evaluation and management of chronic kidney disease. Kidney Int Suppl. 2013;3:1–150. doi:10.1038/kisup.2012.73

